# How much are households willing to invest in hand hygiene enabling technologies? A randomised pricing experiment in Lusaka, Zambia

**DOI:** 10.1101/2025.10.23.25338632

**Authors:** Katherine Davies, Katayi Mwila-Kazimbaya, Ian Ross, Elisabeth Tadiri, Jenala Chipungu, Robert Dreibelbis

## Abstract

Access to a dedicated handwashing facility (HWF) increases handwashing with soap (HWWS), an effective behaviour for preventing respiratory and diarrheal disease. Achieving universal hand hygiene will require household investment of a HWF, yet the ability and willingness of end-users to meet these costs has received little attention in stated or revealed preference studies. This two-phase voucher-based randomised pricing experiment explored whether households in peri-urban Lusaka, Zambia were willing to invest in a HWF. In Phase 1, three HWFs were tested with 60 households using two of the three HWFs for two weeks (20 households per combination). There was strong preference for a locally manufactured bucket with a tap and metal stand (Kalingalinga bucket; 250 ZMW, ∼US$10), over high-cost (1000 ZMW, ∼US$38) and low-cost (100 ZMW, ∼US$4) pre-manufactured HWFs. In Phase 2, n=160 used the Kalingalinga bucket for two weeks. At the end of the trial period, participants received a 50 ZMW gift (∼US$2) and a randomly assigned discount voucher (20%, 40%, 60%, 80%; 40 households per group), before deciding whether to purchase the HWF. Ninety-eight percent (39/40) of households offered an 80% discount purchased the HWF (effective price: 0 ZMW), compared with 30% (12/40) offered a 20% discount (effective price: 150 ZMW). Predictive modelling estimated that 50% of households would purchase at an effective price of 103 ZMW (∼40% of the retail price). Despite declining the purchase offer, all non-purchasing households reported positive stated willingness-to-pay, suggesting latent demand. Findings highlight the need for financial interventions that make desirable HWFs affordable.

## Introduction

Promotion of handwashing with soap (HWWS) is a cost-effective public health measure important for the control of infectious diseases, including acute respiratory infection (ARI) and diarrhoeal disease (Ross et al., 2023; Wolf et al., 2022). Despite this, global adherence to hand hygiene practices remains low, particularly in low-resource settings (Freeman et al., 2014; Wolf et al., 2019). Household access to a dedicated handwashing facility (HWF) increases the likelihood of handwashing with soap (Luby et al., 2009; Wolf et al., 2019) and is important to address the psychological barriers preventing handwashing behaviour (Ezezika et al., 2023; Caruso et al., 2025). Global monitoring efforts to track progress toward Sustainable Development Goal (SDG) 6 targets use the presence of a fixed or mobile HWF as the standard indicator for hand hygiene. Estimates indicate approximately 73% of households in sub-Saharan Africa lack access to a HWF with both soap and water (WHO/UNICEF Joint Monitoring Programme (JMP), 2024a).

The cost of handwashing materials and the resources needed to maintain a handwashing station, as well as a household’s ability to afford them, are frequently cited as barriers to HWWS globally (Ezezika et al., 2023). A recent analysis estimated the total economic cost to reach universal hand hygiene in the 46 least developed countries was US$ 12.2 – 15.3 billion over a 10-year period, with 13% (roughly US$ 1.5 to 2 billion) allocated for the supply of HWFs (Ross et al., 2021). The costs per household for facilities and supplies includes an initial investment of $17 for a HWF (assumed to be a bucket with spigot lasting 5 years), with an annual expenditure of $17 for soap and $5 for water. These costs are typically borne by households. Market-based programming that works through or supports local markets, are increasingly promoted by bilateral and NGO development organisations to improve water, sanitation and hygiene (WASH) services (Tafesse and Tidwell, 2023). However, the extent to which market-based approaches are appropriate for promoting handwashing facility uptake remains unclear.

The ability and willingness of end-users to meet these costs has received little attention in the literature. Willingness-to-pay (WTP) - defined as the maximum amount a consumer is willing to spend for a good or service - can be measured through either stated or revealed preference (Breidert et al., 2006). Stated preference (or stated WTP) is elicited by asking individuals the value they assign to a good or service without actual payment, while revealed preference (or revealed WTP) is estimated from observed purchasing behaviours, in real-world or experimental settings. Multiple studies have found that demand for preventative health goods is highly sensitive to price (Dupas, 2014a) and studies investigating demand and WTP for sanitation products and services often find low WTP in low-income, community settings (Tidwell et al., 2019; Peletz et al., 2017; Peletz et al., 2021; Peletz et al., 2020). A technical brief by World Vision explored voucher redemption rates for a locally designed HWF at different purchase prices (Tafesse and Tidwell, 2023). However, to our knowledge, there appear to be few, if any, stated or revealed preference studies in the published literature to elicit WTP for HWFs. This two-phased voucher-based randomised pricing experiment aimed to explore whether people in peri-urban Lusaka, Zambia were willing to invest their own resources in a HWF.

## Methods

### Study setting and study design

Zambia faces some of the highest levels of poverty and inequality globally. In 2022, an estimated 64% of the population – around 12.6 million people – lived on less than US$ 2.15 per day (Betran et al., 2025). Lusaka is the capital and largest city in Zambia, with a population of over two million. Nearly 62% of Lusaka’s population live in the city’s unplanned and informal settlements (peri-urban areas) (Chiwele et al., 2022). This study was conducted in George and Matero, two peri-urban communities located in Lusaka, Zambia. These communities are characterised by densely populated and informal housing and face recurring cholera outbreaks and other public health challenges linked with inadequate water, sanitation, and hygiene (WASH) services (Hubbard et al., 2020). Approximately 71% of Zambia’s urban population are estimated to lack access to a household HWF with soap and water (WHO/UNICEF Joint Monitoring Programme (JMP), 2024b). These communities were conveniently selected based on existing partnerships.

This study was completed in two phases. Phase 1 of the study explored if end-users were willing to invest in one of three pre-determined HWF designs, and Phase 2 aimed to quantify how much of their own resources end-users were willing to invest in the top-ranking HWF from Phase 1. Below, we present the methods and results for each phase sequentially with a combined discussion.

### Ethical Considerations

Ethical approval for Phase 1 and Phase 2 of the study was gained from London School of Hygiene and Tropical Medicine Research Ethics Committee (Refs: 29842 for Phase 1 and 31268 for Phase 2), and the University of Zambia Biomedical Research Ethics Committee (Refs: UNZABREC 4329-2023 for Phase 1 and UNZABREC 5775-2024 for Phase 2).

Prior to study activities, community sensitization meetings were held with local representatives, who communicated the research through established community platforms. An information sheet was read out by the enumerator to a potential participant outlining the purpose and procedures of the study. Informed consent was obtained from all participants and was confirmed by a thumb-print or written signature, depending on literacy status. Where the individual consenting was not literate, a literate impartial witness signed to confirm that the participant understood and consented to the study.

### Data Management

Data from both phases of the study, including household characteristics and purchasing decisions, were captured using electronic data entry forms built using Open Data Kit (ODK) (Hartung et al., 2010) and administered on android tablets. Primary respondents were the head of household or a nominated adult (18+) household member present at the time of data collection. Data from the tablets were uploaded daily onto a secure, cloud-based server. A team of six research assistants conducted the randomised pricing experiments and data collection. Research assistants were fluent in local languages (Nyanja and Bemba) and had previous experience collecting WASH data and had completed training on data collection procedures and ethical safeguarding (informed consent and data protection).

## Phase 1: Pilot Randomised Pricing Experiment

### Methods - Phase 1

#### Sampling Procedures

Our pilot experiment was embedded within a qualitative study exploring use and acceptability of various handwashing facilities (manuscript in preparation). Sample sizes reflected the estimated maximum enrolment required to reach anticipated diversity between key target end-users for the qualitative study. A purposive sample of 60 households was selected: 18 large households (5+ members, with at least one child under 5), 18 small households (2-4 members), 12 households with members aged 65 or older, and 12 households with members with a disability. These categories informed qualitative sampling only; results from the randomised pricing experiment are not stratified by household type. Community Health Workers (CHWs) familiar with the study communities assisted in identifying households with members aged 65+ or with a member with a disability. Households of varying size were selected using a pre-determined sampling interval across different zones within each community to ensure variation in household characteristics and location.

Inclusion and exclusion criteria were applied at the household level. Households with at least one adult (aged 18 or older) who could consent to the study on behalf of all members of the household were included. Households comprised of a single individual, or households with no adult to provide consent were excluded. Households that already owned a similar HWF were also excluded.

#### Data Collection

Households were recruited and data were collected between 2 May and 10 July 2024. Three HWFs were selected for testing based on a previous ranking exercise of HWFs in the same communities (Mwila-Kazimbaya et al., 2025): the locally manufactured Kalingalinga bucket, the low-cost premanufactured SATO Tap and the relatively high-cost premanufactured Happy Tap (Table 1) (mWater, 2021; Revell and Huynh, 2018). Households were randomly assigned two of the three HWFs, each for a two-week period, in varying sequences (six possible combinations), for a four-week total period. Assignment of HWFs was proportionally balanced across the four household groups and the two communities. New HWFs were provided to all households.

**Table 1.** Summary of handwashing facilines included in the study.

| Name | Approximate Retail Price* | Description |
| --- | --- | --- |
| **Kalingalinga Bucket** | ZMW 250 / 10 USD | A locally manufactured, hand-operated HWF consisting of a 20L bucket with a tap, a small water collection basin, and a supporting frame. It is widely used and well-known in the local setting. |
| **SATO Tap** | ZMW 100 / 4 USD | A low-cost, premanufactured HWF featuring a tap mechanism that attaches to a 0.5-2L water bottle and dispenses water when pressed. Small and portable, allowing for easy relocation. The device is not manufactured locally and is not widely available in Zambia. |
| **Happy Tap** | ZMW 1000 or 38 USD | A high-cost, premanufactured, portable handwashing sink with an 18L water storage tank, a tap, and a catchment tray with drainage. Water is dispensed via a lever that can be operated using the elbow. The device is not manufactured locally and is not widely available in Zambia. |

\*Based on manufacturers or distributor's cost price, plus shipping for HWFs manufactured outside Zambia.

At the end of the four-week period, respondents were asked to state which HWF they preferred using overall, before prices were disclosed. Households were then given a gift of 300 Zambian Kwacha (ZMW), (approximately US$10 in March 2024) and asked to select one of three identical, sealed envelopes containing a voucher for 25%, 50% or 75% off the estimated retail price.

Households could choose to use the gift and discount voucher to purchase one of the two allocated HWFs (Table 2) or to keep the full monetary gift. The gift money was a fungible resource useable for any purpose, while the discount was restricted to HWF purchases. The gift provided participants with liquidity to facilitate the purchasing decision. Households were not informed of the monetary gift at the time of informed consent and were instead advised that options for keeping or purchasing the handwashing facilities would be discussed at the end of the study. Respondents were given up to 10 minutes to allow consultation with other household members. The price reduction was communicated to respondents in absolute value, e.g., “the full price of SATO Tap is ZMW 100, but you can buy it for ZMW 50 with your 50% off discount”.

**Table 2.** Cost and effective price of each handwashing facility based on discount vouchers and 300 ZMW gir.

| Discount Voucher | Happy Tap |  | SATO Tap |  | Kalingalinga Bucket |  |
| --- | --- | --- | --- | --- | --- | --- |
|  | Cost (ZMW) | Effective Price (ZMW)* | Cost (ZMW) | Effective Price (ZMW)* | Cost (ZMW) | Effective Price (ZMW)* |
| **Full Retail Price** | 1000 | - | 100 | - | 250 | - |
| **25% off** | 750 | +450 | 75 | -225 | 187.50 | -112.5 |
| **50% off** | 500 | +200 | 50 | -250 | 125 | -175 |
| **75% off** | 250 | -50 | 25 | -275 | 62.50 | -237.5 |

\*Effective price calculated as the discounted price minus the 300 ZMW gift.

The “effective price” of a HWF was calculated as the out-of-pocket payment a participant would have to contribute to purchase a HWF after the gift money and the discount voucher were combined. Most combinations of facility and voucher amount resulted in negative effective price values, with only the 25% or 50% off vouchers for the Happy Tap resulting in an additional monetary contribution from the household to supplement the gifted funds (Table 2). This pilot phase was not designed around effective price considerations but sought to determine in which HWF households were most inclined to invest.

#### Data Management and Analysis

Data were exported from ODK then uploaded to R (V4.5.0) for data cleaning and analysis. As this was a small pilot study, only descriptive analyses were used to compare overall preferences in HWF design and pricing experiment outcomes.

### Results – Phase 1

A total of 60 households were enrolled in this pilot study, with 30 households from each community. Most respondents were female (88%; 53/60) and lived in a shared plot (75%; 45/60) (Table A in S1 Appendix). Each of the three HWF combinations were allocated to 20 households (10 per community): Kalingalinga bucket + Happy Tap, Kalingalinga bucket + SATO Tap, and Happy Tap + SATO Tap.

#### Stated preference before price

When respondents were asked which of the two HWFs they had tested they preferred using (without reference to price), the locally manufactured Kalingalinga bucket (50%; 30/60) and high-cost premanufactured Happy Tap (45%; 27/60) were favoured over the low-cost premanufactured SATO Tap (5%; 3/60) (Table B in S1 Appendix).

#### Revealed preference

Sixty-seven percent (40/60) of households chose to purchase a HWF rather than keep the gift money, with the Kalingalinga bucket being the most purchased option (53%; 32/60). Purchasing behaviour did not align with stated preferences before price was revealed. Many households who expressed preference for the Happy Tap chose not to purchase any HWF or selected an alternative after the price was revealed (Table B in S1 Appendix). The Happy Tap was only purchased by households who received a 75% discount – effectively covering the full cost – suggesting limited willingness to invest personal resources in this option (Fig 1). When the Kalingalinga bucket was offered alongside another HWF, it was consistently the preferred purchase. No households chose to purchase the SATO Tap over the Kalingalinga bucket, and among those offered the Happy Tap and SATO Tap, most did not purchase either.

**Fig 1.**
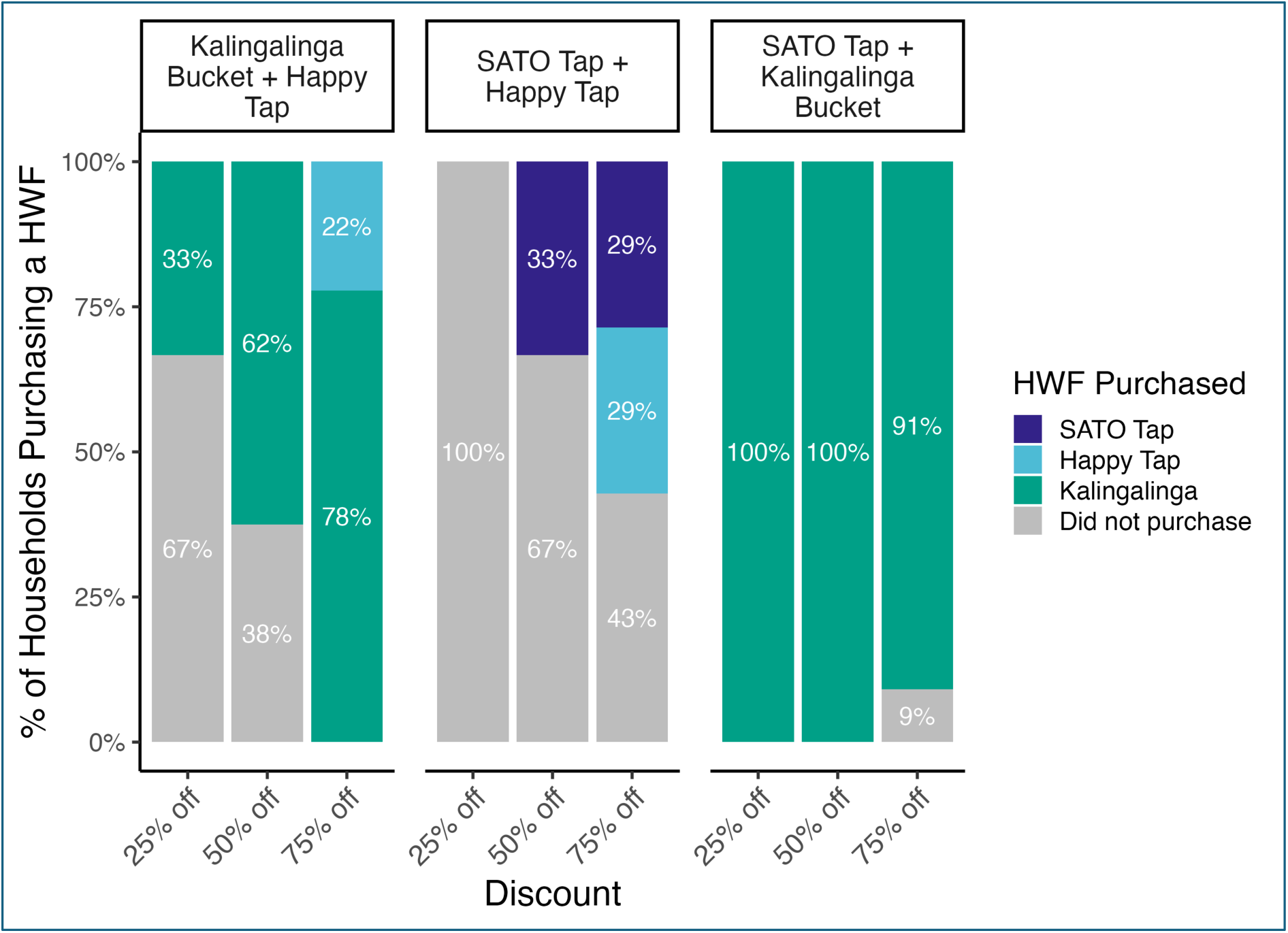
Proportion of households purchasing each HWF by discount category and HWF combination group. *For each combination group, bars show the percentage of households purchasing each of the three HWFs or not purchasing at each voucher level (25%, 50% or 75% off the estimated retail price).*

Although household voucher allocation was random, the resulting distribution was uneven given the small sample size and that each participant was given blinded selection from all 3 vouchers. More households received a 75% discount (45%; 27/60) than a 50% (37%; 22/60) or 25% discount (18%; 11/60). Among households receiving the SATO Tap and Happy tap, voucher allocation was even across discount levels. However, in the other two HWF combinations, a larger proportion of households received higher discounts (Table B in S1 Appendix).

## Phase 2: Randomised Pricing Experiment - Kalingalinga bucket

### Methods – Phase 2

#### Sampling Procedures

To further explore household willingness to pay for HWFs as a function of effective price, a second randomised pricing experiment was completed. A sample of 160 households were recruited for this study (80 households from each community). Given the exploratory nature of the study, the sample size was selected to allow approximation of a demand curve using descriptive analyses, with 40 households per discount level, within the logistical constraints of the study. Assuming a conservative purchase rate of 40%, the planned sample would yield approximately 64 events. With up to five parameters, this provides 12-16 events per parameter, exceeding the recommended minimum for reliable estimation in regression models (Peduzzi et al., 1996).

Households with at least one adult (aged 18 or older) who consented to the study on behalf of all members of the household were included. Households comprised of a single individual, households in which no adult could provide consent, or any household that participated in Phase 1 were excluded. Households that were already using similar HWFs were also excluded. As in Phase 1, households were recruited using a predetermined sampling interval.

#### Data Collection

Households were recruited and data were collected between 27 November to 20 December 2024. Based on the results from Phase 1, this simplified randomised pricing experiment was based only on the Kalingalinga bucket. Households were given a Kalingalinga bucket to use for two-weeks. After two weeks, households were re-visited and given a gift of 50 Zambian Kwacha (ZMW) (approx. USD$2 in Nov 2024). Households were randomly assigned, without replacement, to one of four discount vouchers (1:1:1:1): 20%, 40%, 60% or 80% (40 households per discount group) and were given the option to purchase the Kalingalinga bucket at the discounted price or keep the gift money. Primary respondents were given 5 to 10 minutes to allow consultation with other household members. Purchasing decisions were documented by research assistants in ODK. Table 3 outlines the effective price of the HWF at each discount level.

**Table 3:** Effective price based on discount vouchers and 50 ZMW gift.

| Discount Voucher | Cost to purchase (ZMW) | Effective Price (ZMW) |
| --- | --- | --- |
| 20% | 200 | 150 |
| 40% | 150 | 100 |
| 60% | 100 | 50 |
| 80% | 50 | 0 |

After they had made their decision, we asked respondents to state the maximum amount they would have paid for the Kalingalinga bucket. Stated WTP is generally considered a less reliable reflection of preference than revealed WTP (Fifer et al., 2014), but in this case the participant has just made a revealed preference decision so is in a good position to know their maximum WTP. To ensure internal consistency between stated and revealed preferences, maximum stated WTP values were bounded relative to the discounted price before the gift was applied: respondents who did not purchase a HWF were restricted from stating a maximum WTP above the offered discounted price, while those who did purchase were required to state a WTP greater than or equal to the discounted price at which they purchased.

A questionnaire administered with the primary respondent on the first visit addressed socio-demographic characteristics of the household, asset ownership, WASH access and water insecurity (using the four-item household water insecurity scale, HWISE-4) (Young et al., 2021). Water insecurity was dichotomised, with households scoring 4/12 or above classified as water insecure. A principal component analysis of information on household assets and housing characteristics commonly found in the Demographic and Health Survey (Zambia Statistics Agency, 2019) was conducted to create a relative wealth index. Variables were only included if at least 5% and no more than 95% of households reported ownership to ensure sufficient variation. Twenty-three variables met these criteria and were retained (Table E in S1 Appendix). The first principal component was used to assign each household a score reflective of relative wealth.

#### Data Analysis

Data were exported from ODK then uploaded to R (V4.5.0) for data cleaning and analysis. Data on purchasing decisions were used to estimate a demand curve of the percentage of households purchasing a HWF at each of the four effective price points. Confidence intervals for purchasing proportions at each price point were calculated using exact binomial tests.

Multivariable Poisson regression with robust standard errors was used to examine the association between purchasing decision and effective price. Effective price was modelled as a continuous variable. Hypothesised predictors of purchasing behaviour, including household wealth score, and the age and gender of the primary respondent, as well as study site were adjusted for. Other potential predictors including marital status, income, education, employment, household size and water insecurity were tested but were excluded from the final model.

To estimate the effective price point associated with a 50% predicted probability of purchasing a HWF, the fitted multivariable Poisson regression model was used to generate predicted probabilities across a continuous sequence of effective prices while holding other covariates at median (for continuous) or reference (for categorical) levels.

### Results - Phase 2

A total of 160 households were enrolled in Phase 2 of the study, with 80 households from each of the two study communities (George and Matero). Most respondents were women (96%; 154/160), aged between 35-54 (53%; 84/160), and married (66%; 106/160) (Table C in S1 Appendix). Most households resided on shared plots (76%; 121/160) and had six or more members (50%; 80/160). Discount vouchers were evenly distributed, with 40 households assigned to each discount level, except for 40% and 60% discount groups, which included 39 and 41 households, respectively.

Overall, 63% of all households (101/160) purchased a HWF. Purchasing behaviour varied according to the effective price, with uptake increasing as effective price decreased (Fig 2). At the highest price point of 150 ZMW (20% discount), only 30% (12/40) of households purchased a HWF, compared to 98% (39/40) at the lowest effective price point of 0 ZMW (80% discount). Although confidence intervals overlapped between adjacent price points, the highest and lowest price groups were clearly distinct, indicating an inverse relationship between price and demand consistent with a normal good. Price elasticity of demand appears quite high, especially in the middle of the curve, with an increase in effective price from 50 to 100 ZMW associated with uptake almost halving from 78% to 46%.

**Fig 2.**
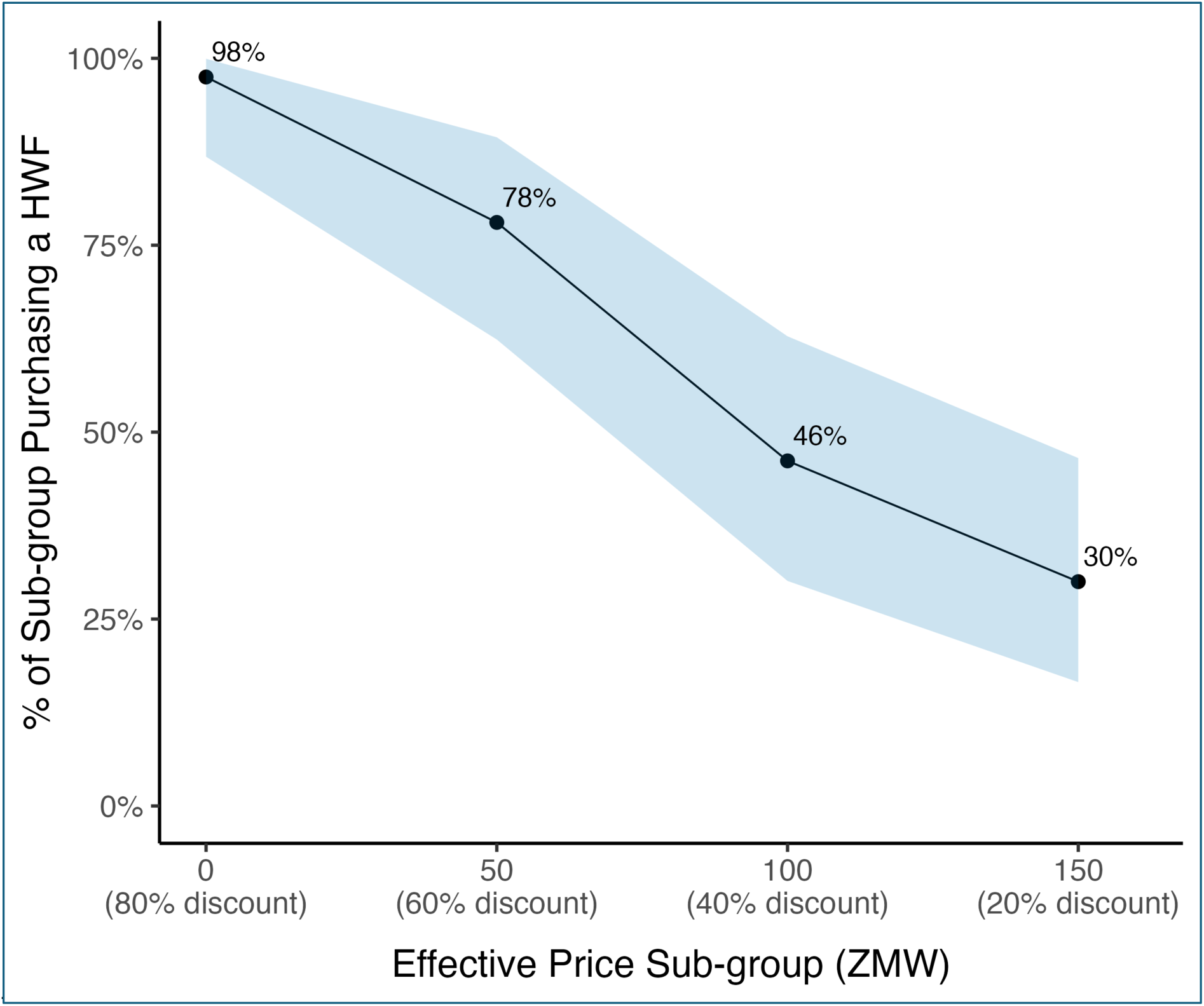
Demand curve showing the proportion of households purchasing a HWF at each effective price point. *The points represent the proportion of households in each effective price sub-group purchasing a HWF, and the shaded ribbon indicates 95% confidence intervals, calculated using exact binomial tests.*

In multivariable Poisson regression, there was a negative association between effective price and purchasing behaviour. After adjustment for respondent age and gender, household wealth, and study site, each 1 ZMW increase in effective price was associated with a 1% decrease in the probability of purchase (adjusted risk ratio = 0.99, 95%CI 0.99-0.99). Full regression results are available in Table D of S1 Appendix. Using the predicted probabilities from the model, an estimated 50% of households would purchase a Kalingalinga bucket at an effective price of 103 ZMW, holding other covariates at median or reference levels (Fig A in Appendix S1).

Stated WTP was higher among households that chose to purchase the HWF (adopters) compared to those who did not (non-adopters) (Fig 3). The median stated WTP among adopters was 150 ZMW (IQR = 100), compared to 100 ZMW (IQR = 50) among non-adopters. The highest stated WTP among non-adopters was 150 ZMW. Despite declining the purchase offer, all non-adopters expressed a positive valuation (max WTP > 0) for the HWF. Very few households provided a maximum stated WTP at or above the retail price.

**Fig 3.**
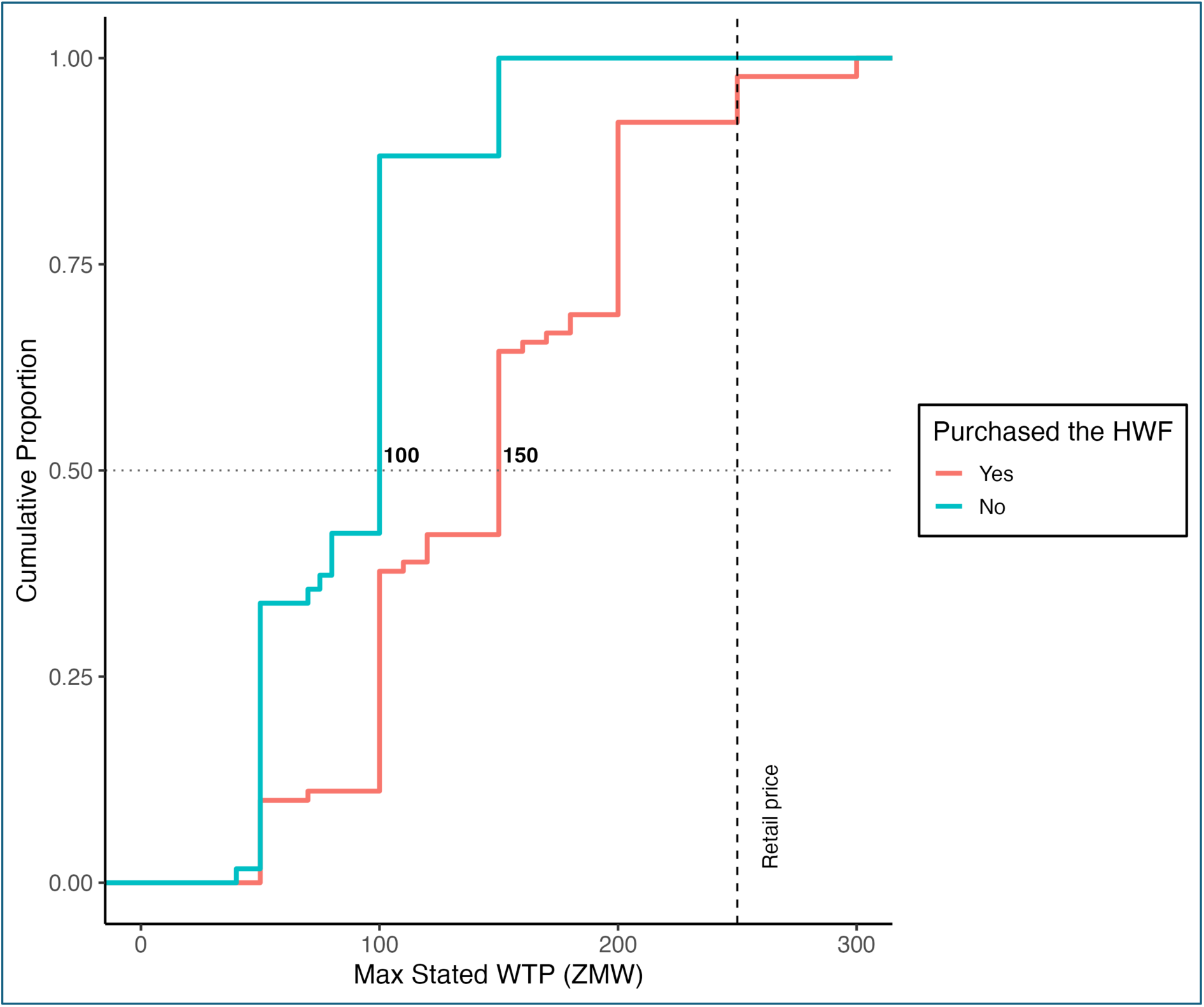
Empirical Cumulative Distribution Function (CDF) plot of maximum stated WTP for the Kalingalinga bucket, by purchasing behaviour. *The figure shows the empirical CDF of the maximum amount households reported they would be willing to pay for the Kalingalinga bucket, stratified by adopters (n = 101) and non-adopters (n = 59). The retail price of the Kalingalinga bucket (250 ZMW) is indicated by a vertical dashed line. Median WTP values for each group are shown as labels alongside the horizonal line at 0.5. Vertical jumps in the lines indicate clustering of responses at certain price points, resulting from discount group allocation and limits on stated maximum WTP according to purchasing behaviour and discount group*.

## Discussion

Achieving universal hand hygiene requires investment, yet the cost of a HWF is typically borne by households (Ross et al., 2021) and little is known about household’s ability or willingness to cover these costs. This study explored purchasing preferences and revealed willingness to pay (WTP) for HWFs in peri-urban Lusaka, Zambia helping to address this critical data gap. In Phase 1, households showed a strong preference for the locally manufactured Kalingalinga bucket (250 ZMW, ∼$10), over higher- and lower-cost premanufactured alternatives. In Phase 2, which focused exclusively on the Kalingalinga bucket, demand was found to be price-sensitive, and regression modelling predicted a 50% purchasing probability at an effective price of 103 ZMW (∼$4). Although retail price was not directly tested, extrapolation suggests uptake would be less than 10% at that price, highlighting a significant affordability gap.

In Phase 1, household preferences were shaped by both design and price. In the absence of cost, households favoured HWFs that resembled a sink (tap and basin), such as the Happy Tap or Kalingalinga bucket, consistent with findings from Tanzania and Zambia (Brial et al., 2023, Mwila-Kazimbaya et al., 2025). With over 40% of households in urban Zambia relying on water sources that are not piped into their or a neighbour’s dwelling (Zambia Statistics Agency, 2019), it is unsurprising that households prefer HWFs that integrate hygiene with broader domestic utility, such as cleaning produce or face-washing. Like multi-purpose prevention technologies in sexual and reproductive health (Karim et al., 2014), HWFs designed for multiple uses may similarly promote greater acceptability, uptake and sustained use. In this study, affordability strongly influenced purchasing, with the lower-cost Kalingalinga bucket favoured over the more expensive Happy Tap. The SATO Tap, despite being cheapest option, was rarely purchased, suggesting design limitations can outweigh affordability in decision-making (Mwila-Kazimbaya et al., 2025). These findings highlight the need to balance affordability with design as well as the urgent need for further innovation in HWF design and production that can meet both the financial resources and household needs of multiple end users.

In Phase 2, the inverse relationship between HWF price and uptake aligns with revealed preferences for sanitation which demonstrate similar price sensitivity (Peletz et al., 2017; Peletz et al., 2021; Peletz et al., 2020). Development economics literature identifies price as a key barrier to the adoption of health-improving technologies, with demand often highly sensitive to small price increases at low price levels, particularly when moving from free (zero) to a small positive price (Spears, 2014). Affordability is frequently cited as a barrier to handwashing in community settings (Ezezika et al., 2023). Closing the affordability gap requires strategies beyond market-driven approaches. Demand-driven interventions such as social marketing, if not paired with mechanisms to alleviate financial barriers, risk widening inequities (Langford and Panter-Brick, 2013; Parvez et al., 2021).

Targeted subsidies or other financing mechanisms can reduce barriers (Dupas, 2014a; Dupas et al., 2016) and evidence from sanitation programming suggests financial support outperforms information or education campaigns in increasing adoption (Gautam et al., 2025). Poverty-targeted subsidies have successfully increased household toilet uptake among low-income households while avoiding market distortions for non-poor households (Nicoletti et al., 2022; Guiteras et al. 2015; Hoo et al., 2022). However, because sanitation has public good characteristics and can generate positive externalities while handwashing infrastructure primarily constitutes a private good, comparisons of subsidy effectiveness across these sectors are limited. Evidence from other preventative health interventions suggests that subsidies can promote sustained adoption. For example, Dupas and colleagues found that one-off subsidies for antimalarial bed nets increased long-term uptake by enabling learning about their benefits, without generating negative price anchoring (Dupas, 2014b). However, they note that such effects may not generalise to other products that differ in durability or familiarity. Therefore, while subsidies have proven effective in sanitation and other health prevention contexts, further research is needed to determine their effectiveness in promoting improved HWF uptake and handwashing behaviour.

Overall uptake of HWFs in our study was high. Over 60% of households purchased a HWF, substantially higher than uptake observed in a randomized pricing experiment for HWFs at WASH Business Centres in Ethiopia (Tafesse and Tidwell, 2023). The high uptake demonstrates the strong value households place on having a HWF with running water at home. Even non-adopters expressed positive valuations (WTP > 0), suggesting latent demand could be unlocked through programming focused on making HWFs more affordable through pricing, financing, and delivery innovations. The high uptake observed may partly reflect features of our study design. The two-week trial period allowed households to test the HWF without knowing they would later be offered the opportunity to purchase it. Diffusion of innovation theory suggests that trialability – the ability to experiment with an innovation on a limited basis - is positively associated with adoption (Rogers, 2003). The opportunity to trial the HWF may therefore have contributed to the high adoption observed. In addition, doorstep delivery of HWFs – rather than voucher redemption used in comparable studies (Peletz et al., 2017; Peletz et al., 2021; Peletz et al., 2020) – likely increased convenience and reduced barriers to uptake. Finally, the short decision window (five minutes), and awareness that field staff were waiting for a response may have introduced social desirability pressures and limited the kind of deliberation that would typically occur in real world purchasing contexts.

This study has several strengths. First, we combined revealed and stated preference methods to elicit WTP. Although stated WTP is considered a less reliable reflection of preference than revealed WTP (Fifer et al., 2014), using both methods together provides a comprehensive understanding of household demand and WTP. Second, the phased approach enabled us to assess willingness to invest in three HWF options, providing insights into household preferences and informing a more focused subsequent phase centred on the most popular technology. We acknowledge the limitations of this study. First, voucher allocation in Phase 1 was uneven, inflating purchase numbers and preventing definitive comparisons across HWF combinations. Second, the experimental setting in Phase 2 is not reflective of real-world conditions. Households used HWFs before purchasing and the product was delivered to their home, creating a more convenient scenario than typical market conditions. Social desirability bias may have also influenced decisions, potentially inflating uptake. In addition, households were only given five minutes to make their decision, which does not reflect the longer deliberation time typical in real-world settings. Third, due to the exploratory nature of the research, sample sizes were small, limiting statistical comparisons and limiting our exploration on determinants of purchasing behaviour. However, we were able to draw meaningful conclusions about the relationship between effective price and purchasing. Finally, findings may not be generalisable beyond populations with similar characteristics, where HWF preferences and valuations could differ

## Conclusion

At current market prices, broad uptake of HWFs above 100 ZMW (∼$4) in peri-urban communities in Lusaka is unlikely. Observed price elasticity of demand suggests that demand-driven strategies to promote handwashing, such as social marketing, risk widening inequities if not paired with mechanisms to alleviate the financial burden on low-income households. Achieving universal hand hygiene will therefore also require interventions that address affordability barriers to ensure all households can access essential infrastructure. Preferences observed in this study reflect trade-offs between design and cost, highlighting the need for innovations that are both affordable and desirable. Moreover, innovations in the promotion and distribution of HWFs could unlock latent demand and expand coverage. Further research should assess how subsidies influence HWF adoption and handwashing behaviour and explore wider household determinants of HWF purchasing behaviour.

## Data Availability

All data produced in the present study are available upon reasonable request to the authors and will be available on LSHTM Data Compass when submitted for publication.

