## Supplementary Materials for "How much are households willing to invest in hand hygiene enabling technologies? A randomised pricing experiment in Lusaka, Zambia"

### Appendix S1

**Table A:** Characteristics of the Phase 1 study sample, by location (N=60)

| Characteristic | Overall<br>N = 60[1] | George<br>n = 30[1] | Matero<br>n = 30[1] |
|---|---|---|---|
| Age (years) | | | |
| *18–34* | 18 / 60 (30%) | 10 / 30 (33%) | 8 / 30 (27%) |
| *35–54* | 26 / 60 (43%) | 9 / 30 (30%) | 17 / 30 (57%) |
| *55+* | 16 / 60 (27%) | 11 / 30 (37%) | 5 / 30 (17%) |
| Gender | | | |
| *Male* | 7 / 60 (12%) | 4 / 30 (13%) | 3 / 30 (10%) |
| *Female* | 53 / 60 (88%) | 26 / 30 (87%) | 27 / 30 (90%) |
| Education | | | |
| *Up to Primary* | 32 / 60 (53%) | 22 / 30 (73%) | 10 / 30 (33%) |
| *Secondary* | 19 / 60 (32%) | 5 / 30 (17%) | 14 / 30 (47%) |
| *More than secondary* | 9 / 60 (15%) | 3 / 30 (10%) | 6 / 30 (20%) |
| Employment | | | |
| *Unemployed* | 15 / 60 (25%) | 7 / 30 (23%) | 8 / 30 (27%) |
| *Housewife/Carer* | 6 / 60 (10%) | 2 / 30 (6.7%) | 4 / 30 (13%) |
| *Casual worker/Piece work* | 4 / 60 (6.7%) | 2 / 30 (6.7%) | 2 / 30 (6.7%) |
| *Self-employed/Business* | 29 / 60 (48%) | 18 / 30 (60%) | 11 / 30 (37%) |
| *Other* | 6 / 60 (10%) | 1 / 30 (3.3%) | 5 / 30 (17%) |
| Marital Status | | | |
| *Single* | 11 / 60 (18%) | 5 / 30 (17%) | 6 / 30 (20%) |
| *Married* | 31 / 60 (52%) | 14 / 30 (47%) | 17 / 30 (57%) |
| *Divorced/Separated/Widowed* | 18 / 60 (30%) | 11 / 30 (37%) | 7 / 30 (23%) |
| Live on Shared Plot | | | |
| *Yes* | 45 / 60 (75%) | 21 / 30 (70%) | 24 / 30 (80%) |
| *No* | 15 / 60 (25%) | 9 / 30 (30%) | 6 / 30 (20%) |

| Characteristic | Overall<br>N = 60[1] | George<br>n = 30[1] | Matero<br>n = 30[1] |
|---|---|---|---|
| Household Group | | | |
| *Disabled member* | 12 / 60 (20%) | 6 / 30 (20%) | 6 / 30 (20%) |
| *Elderly member (65+)* | 12 / 60 (20%) | 6 / 30 (20%) | 6 / 30 (20%) |
| *Large (5-9 members)* | 18 / 60 (30%) | 9 / 30 (30%) | 9 / 30 (30%) |
| *Small (3-4 members)* | 18 / 60 (30%) | 9 / 30 (30%) | 9 / 30 (30%) |

[1]n / N (%)

**Table B:** Stated HWF preference, discount voucher allocation, and HWF purchasing decision, by HWF combination group (N=60; n=20 per group).

| Characteristic | Overall<br>N = 60[1] | Kalingalinga Bucket + Happy Tap<br>n = 20[1] | SATO Tap + Happy Tap<br>n = 20[1] | SATO Tap + Kalingalinga Bucket<br>n= 20[1] |
|---|---|---|---|---|
| **Stated HWF Preference** | | | | |
| *Happy Tap* | 27 / 60 (45%) | 9 / 20 (45%) | 18 / 20 (90%) | - |
| *Kalingalinga* | 30 / 60 (50%) | 11 / 20 (55%) | - | 19 / 20 (95%) |
| *SATO Tap* | 3 / 60 (5.0%) | - | 2 / 20 (10%) | 1 / 20 (5.0%) |
| **Discount Voucher** | | | | |
| *25% off* | 11 / 60 (18%) | 3 / 20 (15%) | 7 / 20 (35%) | 1 / 20 (5.0%) |
| *50% off* | 22 / 60 (37%) | 8 / 20 (40%) | 6 / 20 (30%) | 8 / 20 (40%) |
| *75% off* | 27 / 60 (45%) | 9 / 20 (45%) | 7 / 20 (35%) | 11 / 20 (55%) |
| **HWF Purchased** | | | | |
| *Happy Tap* | 4 / 60 (6.7%) | 2 / 20 (10%) | 2 / 20 (10%) | - |
| *Kalingalinga* | 32 / 60 (53%) | 13 / 20 (65%) | - | 19 / 20 (95%) |
| *SATO Tap* | 4 / 60 (6.7%) | - | 4 / 20 (20%) | 0 / 20 (0%) |
| *Did not purchase* | 20 / 60 (33%) | 5 / 20 (25%) | 14 / 20 (70%) | 1 / 20 (5.0%) |

[1]n / N (%)

**Table C:** Characteristics of the Phase 2 study sample and HWF purchasing within each subgroup (N=160).

| **Characteristic** | **Total N = 160** [1] | **Household purchased a HWF**[1] | **Household did not purchase a HWF**[1] |
|---|---|---|---|
| Purchasing Behaviour | | | |
| *Purchased a HWF* | 101 / 160 (63%) | - | - |
| *Did not purchase a HWF* | 59 / 160 (37%) | - | - |
| Discount | | | |
| *20% off* | 40 / 160 (25%) | 12 / 40 (30%) | 28 / 40 (70%) |
| *40% off* | 39 / 160 (24%) | 18 / 39 (46%) | 21 / 39 (54%) |
| *60% off* | 41 / 160 (26%) | 32 / 41 (78%) | 9 / 41 (22%) |
| *80% off* | 40 / 160 (25%) | 39 / 40 (98%) | 1 / 40 (2.5%) |
| Community | | | |
| *George* | 80 / 160 (50%) | 52 / 80 (65%) | 28 / 80 (35%) |
| *Matero* | 80 / 160 (50%) | 49 / 80 (61%) | 31 / 80 (39%) |
| Age of Respondent (years) | | | |
| *18–34* | 63 / 160 (39%) | 35 / 63 (56%) | 28 / 63 (44%) |
| *35–54* | 84 / 160 (53%) | 55 / 84 (66%) | 29 / 84 (35%) |
| *55+* | 13 / 160 (8.1%) | 11 / 13 (85%) | 2 / 13 (15%) |
| Gender of Respondent | | | |
| *Male* | 6 / 160 (3.8%) | 6 / 6 (100%) | 0 / 6 (0%) |
| *Female* | 154 / 160 (96%) | 95 / 154 (62%) | 59 / 154 (38%) |
| Marital Status of Respondent | | | |
| *Single* | 22 / 160 (14%) | 12 / 22 (55%) | 10 / 22 (45%) |
| *Married* | 106 / 160 (66%) | 71 / 106 (67%) | 35 / 106 (33%) |
| *Divorced/Separated/Widowed* | 32 / 160 (20%) | 18 / 32 (56%) | 14 / 32 (44%) |
| Monthly Income (ZMW)* | | | |
| *0-1000* | 27 / 146 (18%) | 18 / 27 (67%) | 9 / 27 (33%) |
| *1001-2500* | 60 / 146 (41%) | 43 / 60 (72%) | 17 / 60 (28%) |
| *2501-5000* | 43 / 146 (29%) | 24 / 43 (56%) | 19 / 43 (44%) |

| Characteristic | Total<br>N = 160 [1] | Household purchased a HWF[1] | Household did not purchase a HWF[1] |
|---|---|---|---|
| *5001+* | 16 / 146 (11%) | 7 / 16 (44%) | 9 / 16 (56%) |
| Education (Head of Household) | | | |
| *Up to Primary* | 39 / 160 (24%) | 26 / 39 (67%) | 13 / 39 (33%) |
| *Secondary* | 108 / 160 (68%) | 68 / 108 (63%) | 40 / 108 (37%) |
| *More than secondary* | 13 / 160 (8.1%) | 7 / 13 (54%) | 6 / 13 (46%) |
| Employment (Head of Household) ** | | | |
| *Unemployed or Retired* | 19 / 159 (8.2%) | 12 / 19 (63%) | 7 / 19 (37%) |
| *Casual worker/piece work* | 29 / 159 (18%) | 21 / 29 (72%) | 8 / 29 (28%) |
| *Self-employed/Business* | 80 / 159 (50%) | 52 / 80 (65%) | 28 / 80 (35%) |
| *Formal Employee* | 31 / 159 (19%) | 15 / 31 (48%) | 16 / 31 (52%) |
| Household Water Insecurity | | | |
| *Water Insecure (≥4)* | 101 / 160 (63%) | 65 / 101 (64%) | 36 / 101 (36%) |
| *Water secure (<4)* | 59 / 160 (37%) | 36 / 59 (61%) | 23 / 59 (39%) |
| Primary Drinking Water Source | | | |
| *Piped into dwelling* | 6 / 160 (3.8%) | 6 / 6 (100%) | 0 / 6 (0%) |
| *Piped into compound, yard or plot* | 87 / 160 (54%) | 56 / 87 (64%) | 31 / 87 (36%) |
| *Piped to neighbour* | 17 / 160 (11%) | 8 / 17 (47%) | 9 / 17 (53%) |
| *Public tap/standpipe* | 42 / 160 (26%) | 26 / 42 (62%) | 16 / 42 (38%) |
| *Borehole or tubewell* | 5 / 160 (3.1%) | 4 / 5 (80%) | 1 / 5 (20%) |
| *Sachet water* | 3 / 160 (1.9%) | 1 / 3 (33%) | 2 / 3 (67%) |
| Household Size | | | |
| *1–3* | 19 / 160 (12%) | 12 / 19 (63%) | 7 / 19 (37%) |
| *4-5* | 61 / 160 (38%) | 37 / 61 (61%) | 24 / 61 (39%) |
| *6+* | 80 / 160 (50%) | 52 / 80 (65%) | 28 / 80 (35%) |
| Live on Shared Plot | | | |
| *Yes* | 121 / 160 (76%) | 75 / 121 (62%) | 46 / 121 (38%) |
| *No* | 39 / 160 (24%) | 26 / 39 (67%) | 13 / 39 (33%) |
| Wealth (Asset Score) | -0.13 (1.90) | -0.17 (1.60) | -0.12 (2.60) |

| Characteristic | Total<br>N = 160 [1] | Household purchased a HWF[1] | Household did not purchase a HWF[1] |
|---|---|---|---|

[1] n / N (%); Median (IQR)

*Missing data for 14 households

**Missing data for 1 household

**Table D:** Full regression results: adjusted estimates of the risk ratios for purchasing behaviour estimated by Poisson regression with robust standard errors (N=160).

| Characteristic | Adjusted RR† | 95% CI | P-value* |
|---|---|---|---|
| Effective Price | 0.99 | 0.99, 0.99 | <0.001 |
| Age (years) | 1.01 | 1.00, 1.02 | 0.007 |
| Gender | | | |
| Male | 1.00 | - | - |
| Female | 0.95 | 0.77, 1.18 | 0.664 |
| Wealth | 0.98 | 0.92,1.03 | 0.369 |
| Study Site | | | |
| George | 1.00 | - | - |
| Matero | 0.93 | 0.76, 1.13 | 0.436 |

Abbreviations: CI = Confidence Interval, RR = Risk Ratio

† Adjusted for age and gender of primary respondent, household wealth and study site. Compared to the crude RR of 0.99 (95%CI 0.99-0.99).

*From the Wald Test

**Table E:** Output of the principal component analysis displaying the proportion of variance of each variable explained by the first component.

| Variable | Proportion of variance |
|---|---|
| Owns a house | 0.22 |
| Owns land | 0.12 |
| Owns Livestock: chickens | 0.11 |
| *Asset ownership* | |
| Bank account | -0.04 |
| Bicycle | 0.14 |
| Car | 0.24 |
| Computer | 0.17 |
| Electricity | 0.25 |
| Internet | 0.12 |
| Microwave | 0.22 |
| Radio | 0.24 |
| Refrigerator | 0.28 |
| Sofa | 0.16 |
| Table | 0.24 |
| TV | 0.32 |
| Watch | 0.18 |
| *Household characteristics* | |
| Energy for cooking: Charcoal | -0.22 |
| Main floor material: Cement | -0.31 |
| Main floor material: Ceramic tiles | 0.17 |
| Main roof material: Asbestos | -0.17 |
| Main roof material: Metal/Iron sheets | 0.18 |
| Main wall material: Cement | -0.07 |
| Main wall material: Cement blocks | 0.06 |

N.B. The first principal component accounts for 16% of the variance, with an eigenvalue of 3.57.

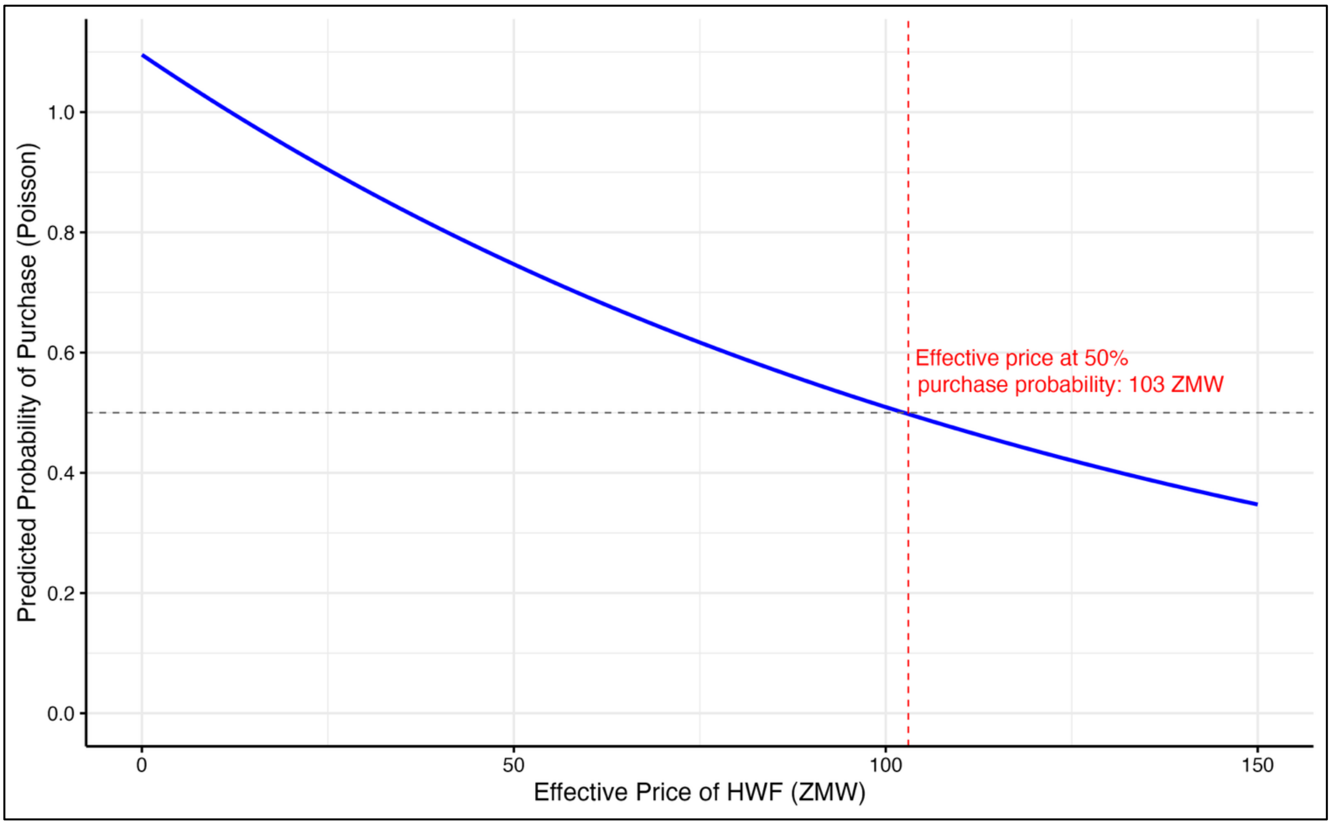


**Fig A. Predicted Probability of Purchase by Effective Price (ZMW).** *Dashed lines indicate the estimated effective price where 50% of households would purchase a HWF.*
